# Effect of and Interventions in Prevention and Management of Maternal Anemia in the Advent of COVID-19

**DOI:** 10.1101/2024.02.18.24302492

**Authors:** John K. Muthuka, Diana Fondo, Francis Wambura, Japheth Mativo, Oluoch J. Kelly, Rosemary Nabaweesi

## Abstract

**Background:** There were many unknowns for pregnant women during the COVID-19 pandemic. Most of these could have been silent however lethal and anemic conditions could escalate the worsening of pregnancy outcomes. Existing evidence indicate that, array of factors is associated with the ability of compromising maternal anemia, some directly and others indirectly.

**Objective:** This review aimed at ascertaining the pooled effect of several anemia interventions. Specifically, the aim of this study was to establish if pregnancy status is associated with COVID-19 severity characterized by a cytokine storm.

**Methods:** We searched the Google Scholar, PubMed, Scopus, Web of Science, and Embase databases to studies suitable for inclusion in this meta-analysis. Studies examining women of reproductive age on any maternal anemia intervention were included. The risk of bias was assessed using the Cochrane risk of bias tool. Review Manager 5.4.1 was used to calculate rate ratios (RRs) with 95% CIs, which were depicted using forest plots. Quantitative variables were summarized in total numbers and percentages. The effect on prevention, control, management and or treatment of anemia was calculated and compared between the intervention and the comparator arms. Heterogeneity was evaluated with the Cochran Q statistic and Higgins test.

**Results:** A total of 11 articles including data for 6,129 were included. With sensitivity analysis, the interventions had a utility of 39% on maternal anemia prevention and management (random effects model RR 0.61, 95% CI 0.43, 0.87; *P* = 0.006) (χ^2^6=286.98, P<.00001; I^2^=97%). All the interventions against maternal anemia showed an effect of 17% (fixed-effect model RR 0.83, 95% CI 0.79-0.88; P<.00001) (χ^2^4=2.93, P=0.57; I^2^=0%). Education to pregnant women showed a 28% effect (RR 0.72 95% CI 0.58, 0.89), medicinal administration 19% (RR 0.81 95% CI 0.73, 0.90), iron supplementation 17% (RR 0.83 95% CI 0.75, 0.92) and I.V Ferric Carboxy-maltose 15% (RR 0.85 95% CI 0.74, 0.97) (I^2^ = 0%). Interventions in African region had a higher (16%) and significant effect compared to other regions (fixed-effects model RR 0.84, 95% CI 0.79-0.89; P<.001) (χ^2^5=176.53, P<.00001; I^2^=97%). Multiple center studies had a significant predictive effect (16%) compared to single center studies (fixed-effects model RR 0.84, 95% CI 0.79-0.89; P<.00001)(χ^2^5=176.53, P<.00001; I^2^=97%). The year 2020 recorded the highest effect of maternal anemia interventions at 28% (random-effects model RR 0.72, 95% CI 0.67-0.78; P<.00001) (χ^2^3=167.34, P<.00001; I^2^=98%)

**Conclusion:** In the advent of COVID-19, maternal anemia interventions were compromised demonstrated by a low effectiveness trend from the year 2020 to the year 2022. During this period, even the most effective and recommended interventions against maternal anemia were somehow affected.

## Introduction

Anemia is a condition in which the number of red blood cells or the hemoglobin concentration within them is lower than normal. Maternal anemia is the percentage of women with Hb levels less than 12 g/dL[1], [2]. Prevalence of anemia in women has been found to be correlated with gross domestic product (GDP) per capita while projections predict a 10% of global GDP decline due to COVID-19 with findings that, the availability of nutritious foods in particular is affected by COVID-19 measures[3].

About one in four women conceive with inadequate or absent iron stores with the levels of serum ferritin below 30 mg/l, and up to 90% have iron stores of below 500 mg, or with serum ferritin below 70 mg/l [4]. These levels are insufficient to meet the increased iron needs during pregnancy, delivery, and postpartum. Moderate to severe anemia in pregnancy especially at 28 weeks and above contributes to 23% maternal mortality globally [5]. It is associated with parasitic diseases such as malaria and worm infestations, acute or chronic illnesses such as sickle cell anemia, tuberculosis, HIV infection, and different macronutrient disorders[6], [7].

Anemia is associated with increased morbidity and mortality in women and children[8], poor birth outcomes [9], decreased work productivity in adults[10], and impaired cognitive and behavioral development in children. Preschool children (PSC) and women of reproductive age (WRA) are particularly affected[11].

The World Health Assembly set six targets to be accomplished by the year 2025. Among the targets is a 50% reduction of anemia in women of reproductive age through several strategies such as food fortification with iron, folic acid, and other micronutrients, distribution of iron-containing supplements, control of infections and malaria [12].

Globally, the COVID-19 pandemic has had devastating effects on health care delivery systems for people of all ages, but pregnant women face particular challenges [13]. Reports show that the pandemic is making it increasingly challenging to provide adequate maternity care worldwide[13]. Even the movement of people seeking to access health care services has been restricted in many countries to prevent the spread of the virus. The pandemic has led to a complete stoppage of the import and export of many essential commodities among various countries, leading to a shortage of necessary items and affecting healthcare services badly, especially sexual and reproductive health care [14]. The population is advised not to attend hospital unless strictly necessary; this advice seems to apply to all, including healthy pregnant women and even those with complications [13].

There were many unknowns for pregnant women during the COVID-19 pandemic. Most of these could have been silent however lethal and anemic conditions could escalate the worsening of pregnancy outcomes. The possibility to this would be accustomed to intervention efforts being compromised due to the COVID-19 pandemic as it has been common with similar pandemics which have been shown to affect the effect of health interventions in vulnerable populations[15].

The direct and indirect effects of the COVID-19 response on pregnant women, newborn babies, young children, and adolescents are enormous, and possibly this, translates to interventions meant to mitigate the anemic conditions in pregnancy. This review aimed at ascertaining the pooled effect of several anemia interventions. Specifically, the aim of this study was to establish if pregnancy status is associated with COVID-19 severity characterized by a cytokine storm.

## Methods

### Design

All guidelines listed in the PRISMA (Preferred Reporting Items for Systematic Reviews and Meta-Analyses) statement were followed in performing this meta-analysis [16]. For this systematic review and meta-analysis, data were pooled from observational studies, including cohort, case-control, cross-sectional, and similar viable case studies. The study was PROSPERO registered (CRD-CRD42023410657).

### Search Strategy

We performed a simple search in the Google Scholar, PubMed, Scopus, Web of Science, and Embase databases to identify observational studies suitable for inclusion with the following search terms: “maternal anemia” OR “anemic condition” OR “poor hemoglobin levels)” OR “pregnancy anemia” OR “anemia in pregnant women” OR “gestation anemia” AND “treatment” OR “intervention” OR “management” AND “effect” OR “effectiveness” AND “impact” OR “outcome” Studies were restricted to those published in English from December 2019 to August 2022.

### Inclusion and Exclusion Criteria

Inclusion criteria were as follows: (1) studies that examined women within reproductive age and put on any anemia prevention program or intervention, either anemic or non-anemic according to World Health Organization (WHO) criteria; (2) observational, cross-sectional, prospective, or retrospective studies; (3) studies that compared intervention approaches with control or comparator approaches; (4) studies evaluating the effects of different interventions among pregnant in the advent of COVID-19.

Exclusion criteria were as follows: (1) unrelated, duplicated, and missing information answering our research question; (2) non-English-language studies; (3) case reports/series; (4) reviews; (5) editorials; (6) studies lacking a full text (unavailable or not yet published); (7) articles without a DOI; and (8) studies with small sample sizes (<50 patients) because of low statistical power.

Notably, we included preliminary findings published as preprints given that the phenomenon in question remains very grey in the public domain and thus we presumed inclusion of such reports would be of value in converging relevant data and information.

### Data Extraction

Both adjusted and non-adjusted data among pregnant on interventions versus pregnant women on comparators arm were extracted to identify the most relevant confounding factors to be used in the analysis by subsequent pooling. Two reviewers (JM and MK) scanned study titles and abstracts obtained via an initial database search and included relevant articles in a secondary pool. Next, two independent reviewers (FM and KO) evaluated the full texts of these articles to determine whether they met the study inclusion criteria. Any disputes were resolved by discussion and negotiation with a fourth reviewer (NM). Only studies agreed upon by all reviewers were included in the final analysis.

The following data were obtained from all studies: title, first author, data collection year, region, sample size, study design, study setting (single or multicenter), intervention type, and the effect associated with each intervention approach. The analysis was then performed to determine whether the intervention group was more likely to portray a better effect on maternal anemia mitigation, treatment or management by ascertaining the end result indicators such as hemoglobin level, and further, sensitivity and sub-group analysis was used

### Risk of Bias (Quality) Assessment

To assess the quality of the included randomized controlled trials, the risk of bias was assessed using the Cochrane risk of bias tool, and the RoB 2 tool (7.0) [17]. The included randomized controlled trials will be assessed in the following domains: risk of bias arising from the randomization process, risk of bias due to deviations from the intended interventions, risk of bias due to missing outcome data, risk of bias in measurement of the outcome, and risk of bias in selection of the reported result, will be followed by the assessment of the overall risk of bias. On the other end, the National Institutes of Health tool for observational and cross-sectional studies [18] was used for methodological quality assessment. Two to three reviewers independently assessed the quality of the studies, and the scores were added to the data extraction form before inclusion in the analysis to reduce the risk of bias. To evaluate the risk of bias, the reviewers rated each of the 14 items into qualitative variables: yes, no, or not applicable. An overall score was calculated by adding the scores of all items with yes=1 and no or not applicable=0. A score was given for every paper, resulting in a classification of poor (score 0-5), fair (score 6-9), or good (score 10-14). Data were checked by reviewers who did not perform the data extraction or each reviewer was assigned an article that they had not extracted data from in previous steps; however, in rare instances, some reviewers extracted data and performed the quality assessment for the same article.

### Statistical Analyses

Review Manager 5.4.1 was used to calculate rate ratios (RRs) with 95% CIs, which are depicted using forest plots. Quantitative variables were summarized in terms total numbers and percentages. The RR not favoring the intervention arm (favoring the control/comparator arm). The effect on prevention, control, management and or treatment of anemia was calculated and compared between the intervention and the comparator/control arms. Heterogeneity was evaluated with the Cochran Q statistic and Higgins test. The Higgins test uses a fixed-effects model when the heterogeneity is <50% and a random-effects model when the heterogeneity is >50%. When heterogeneity was detected, a sensitivity adjustment was made to determine its source. This procedure was performed by leaving a study out of the analysis one at a time, with the fixed-effects model applied after excluding heterogeneity. Subgroup, cumulative analyses, and meta-regression were used to test whether or not the results were consistent and to investigate the effect of confounders on the outcome (anemia control or anemic condition mitigation indicators) and elucidate the best predictors of the outcome. Publication bias was evaluated using the Cochrane Risk of Bias tool.

## Results

### Included Articles and Quality Assessment

The initial search of international databases using the keywords described above yielded 248 articles. After excluding 110 duplicate articles, 138 articles remained. When article titles and abstracts were evaluated for appropriateness, 33 articles ultimately met the inclusion criteria. In addition, 22 articles not meeting the inclusion criteria were excluded after full-text review. A total of 11 articles met the inclusion criteria[19], [20], [29], [21]–[28]. Multimedia Appendix 1 shows the PRISMA flow diagram of the study selection procedure.

**Multimedia Appendix 1:**
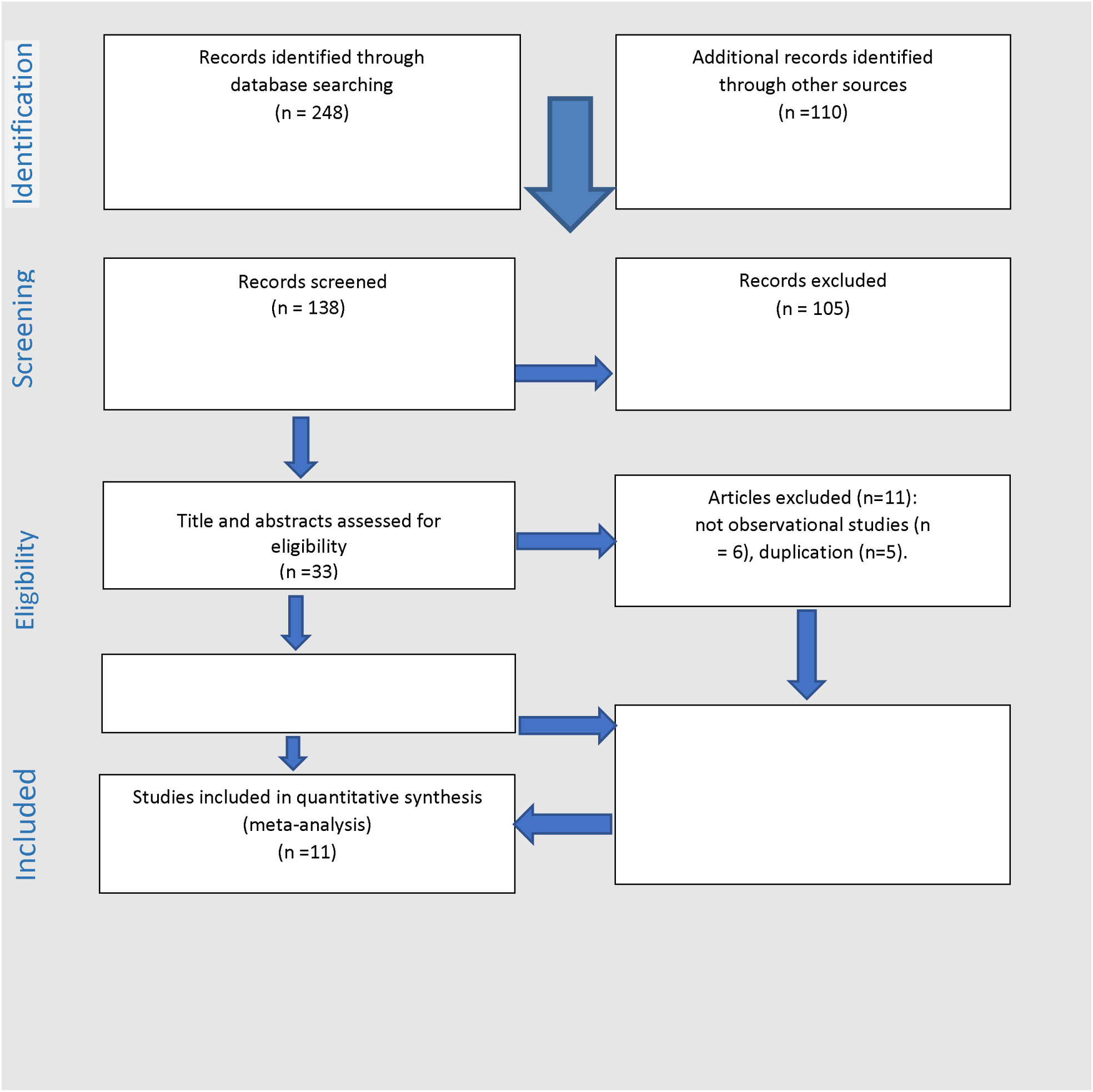
The PRISMA flow diagram of the study selection procedure Identification of studies via database and registers.

### Features of the Included Studies

The 11 included studies provided data for 6,129 pregnant women in the advent of COVID-19 pandemic[19], [20], [29], [21]–[28] *(Table 1),*with 3,591 (59%) pregnant women in maternal anemia intervention group and 2538 (41%) pregnant women in comparator or control group. Among the pregnant women in the intervention group, 53.4% (1921/3591) had the effect of the intervention reported, with similar proportion 53.1% (1350/2538) among the pregnant women in the comparator group. The cumulative effect of both the intervention and the comparator on maternal anemia from all studies ranged from 23% to 81% (average 56%).

**Table 1.**
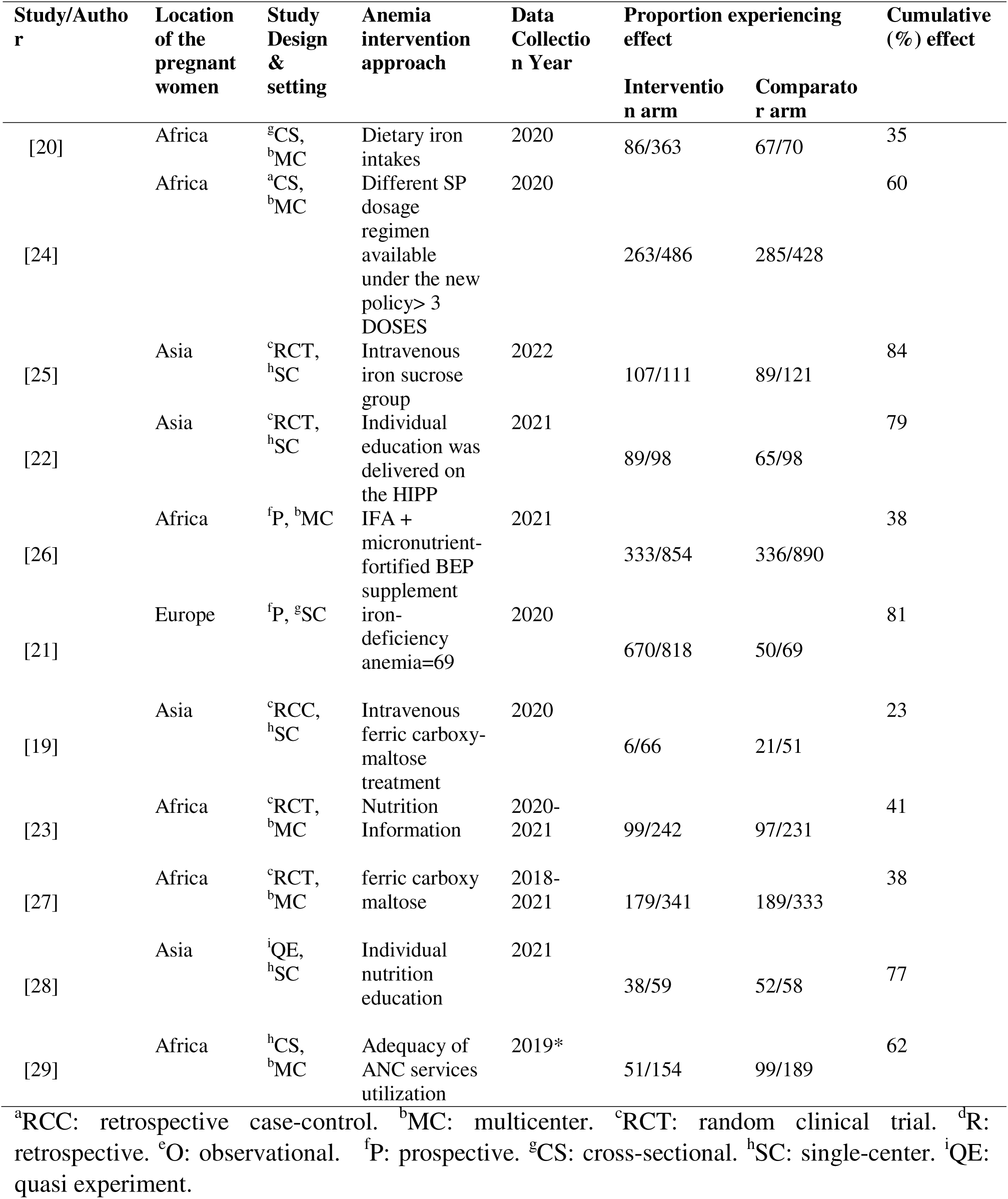
Features of the studies included in the meta-analysis.

The main outcome of this meta-analysis was the pooled effect of interventions on maternal anemia, which was indicated by different outcomes perceived to be due to the specific form of intervention. The key parameter used for assessment of improved maternal anemia post-intervention was the increased levels of hemoglobin alongside other parameters. The study designs included RCTs (n=4, 2 multicenter and 2 single-center studies), cross sectional (n=3, all multicenter), prospective (n=2, 1 multicenter and 1 single-center studies), retrospective case-control (n=1, single-center study) and quasi experiment (n=1, single center). A summary of the studies included in the meta-analysis is provided in *Table 1*.

We assessed the quality of the included observational studies based on a modified version of the Newcastle-Ottawa Scale (NOS), which consists of 8 items with 3 subscales, and the total maximum score of these 3 subsets is 9. We considered a study that scored ≥7 to be a high-quality study since a standard criterion for what constitutes a high-quality study has not yet been universally established. The 11 studies assessed generated a mean value of 6.7, indicating that the overall quality was moderate (NOS score range 5-8), as detailed in *Table 2*.

**Table 2.**
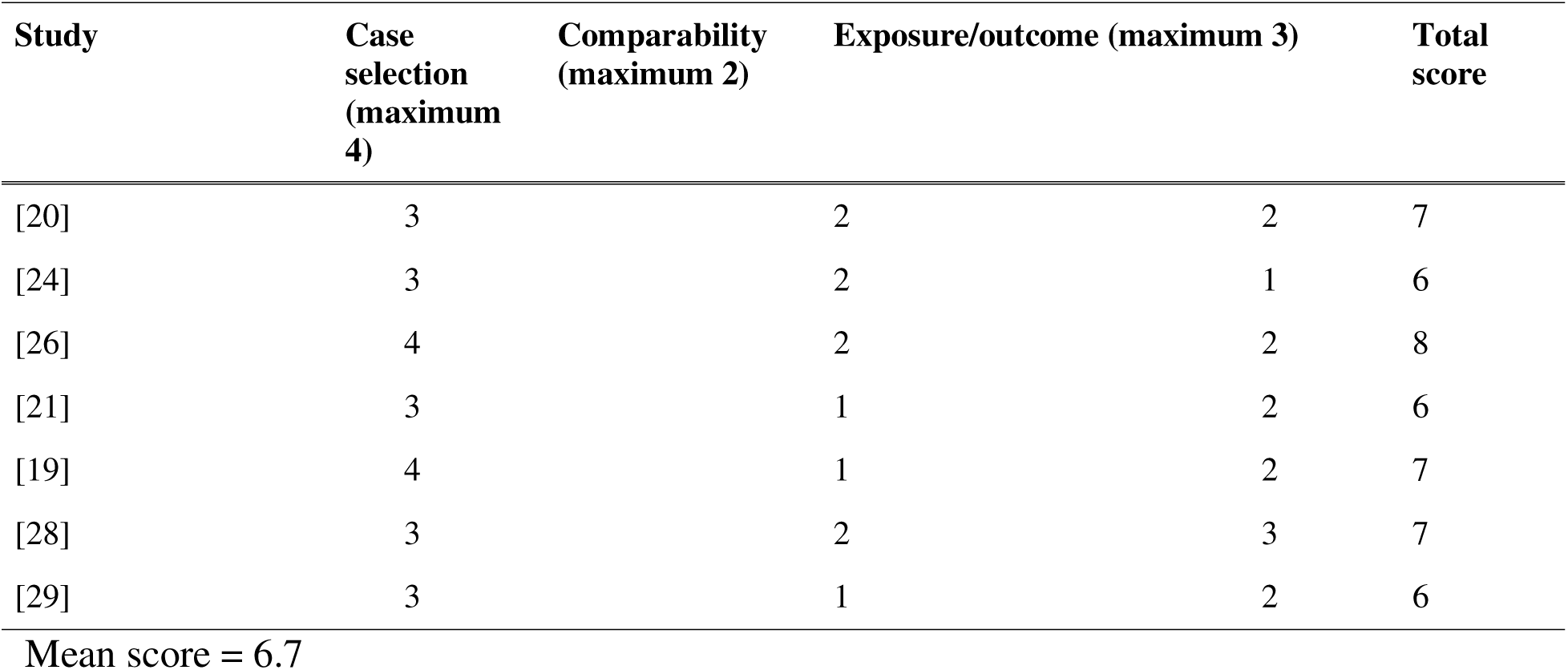
Newcastle-Ottawa scale for quality assessment and risk of bias.

### The pooled effect of interventions on prevention and management of maternal anemia

The meta-analysis revealed a non-significant effect of the interventions on the on prevention and management of maternal anemia indicated by stabilized hemoglobin levels and other parameters (random-effects model RR 0.79, 95% CI 0.61-1.02; P=.07) (χ^2^10=286.98, P<.00001; I^2^=97%). Based on the confidence interval, this indicated little knowledge about the effect and this imprecision affected the certainty in the evidence, thus, further information was needed before a more certain conclusion (*Figure 1*). A funnel plot demonstrated asymmetrical shape depicting the presence of publication bias (*Figure 2*).

**Figure 1.**
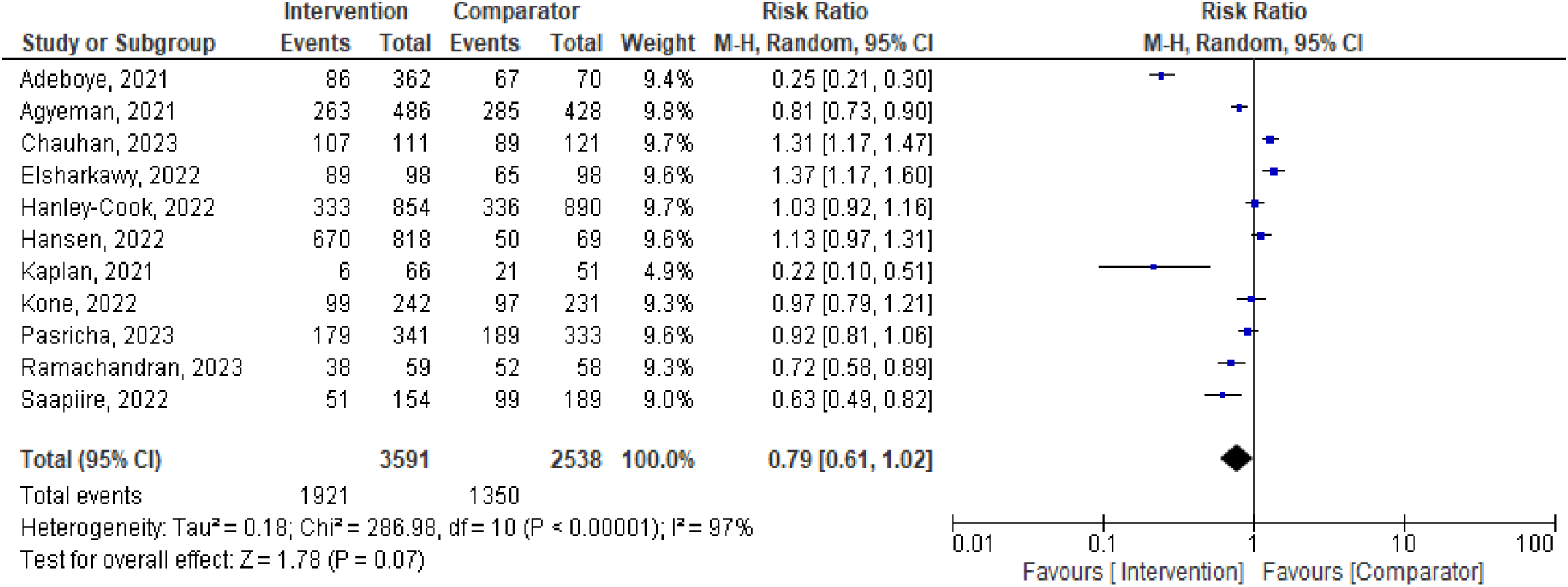
A forest plot of meta-analysis on the effect of maternal anemia interventions

**Figure 2.**
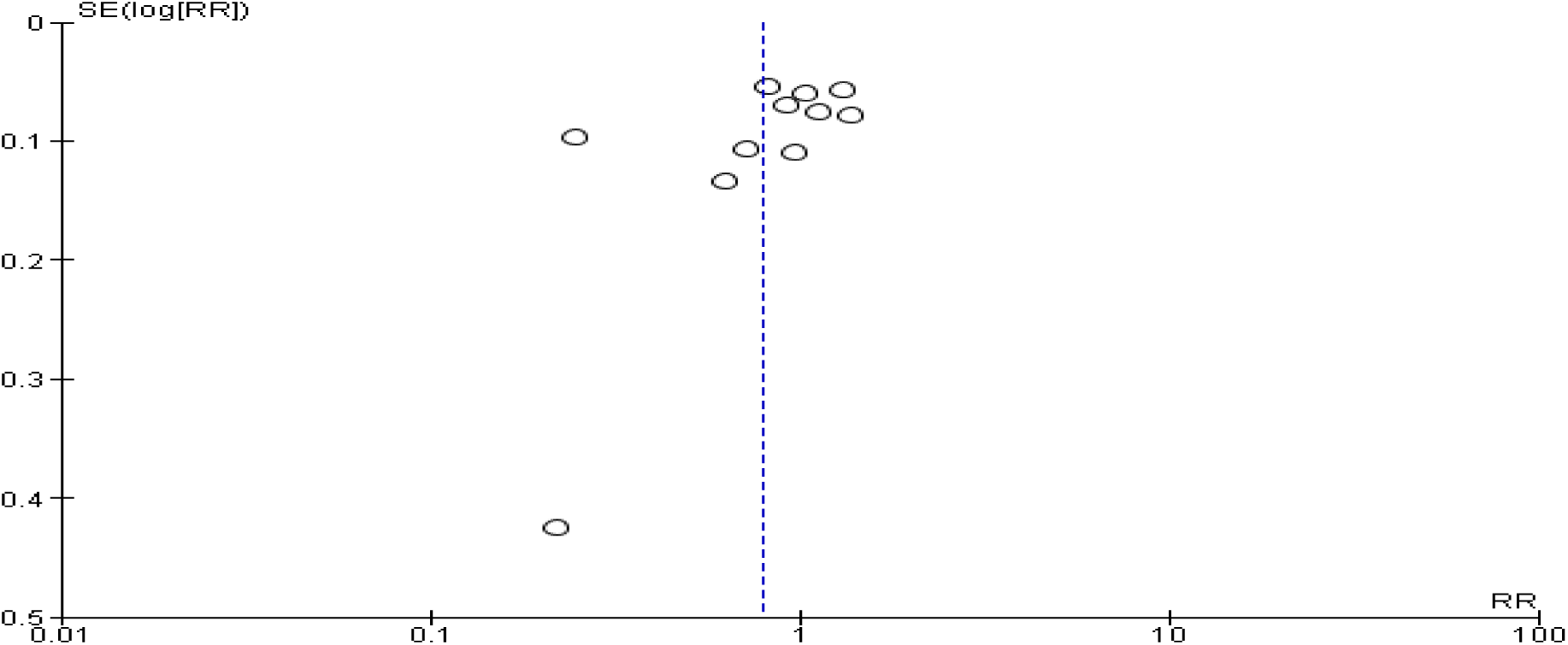
Funnel plot evaluating publication bias. RR: relative risk, SE: standard error

A sensitivity analysis was thus performed to explore the impact of excluding or including studies in the meta-analysis based on sample size, methodological quality, and variance. After removing four studies (n=1,788 pregnant women) [21]–[23], [25] with wider 95% C.I (confidence intervals), a total of 4,341 pregnant women were left for analysis in the remaining studies[19], [20], [24], [26]–[29]. *Figure 3* shows a shift to (random effects model RR 0.61, 95% CI 0.43, 0.87; *P = 0.*006) (χ^2^6=286.98, P<.00001; I^2^=97%) revealing that, the interventions had a utility of 39% on maternal anemia prevention and management in the advent of COVID-19 (Figure 3). The funnel plot evaluating publication bias, revealed considerable heterogeneity between all pooled studies for the updated analysis (*I^2^*=97%, *P<.*00001) (*Figure 4*).

**Figure 3.**
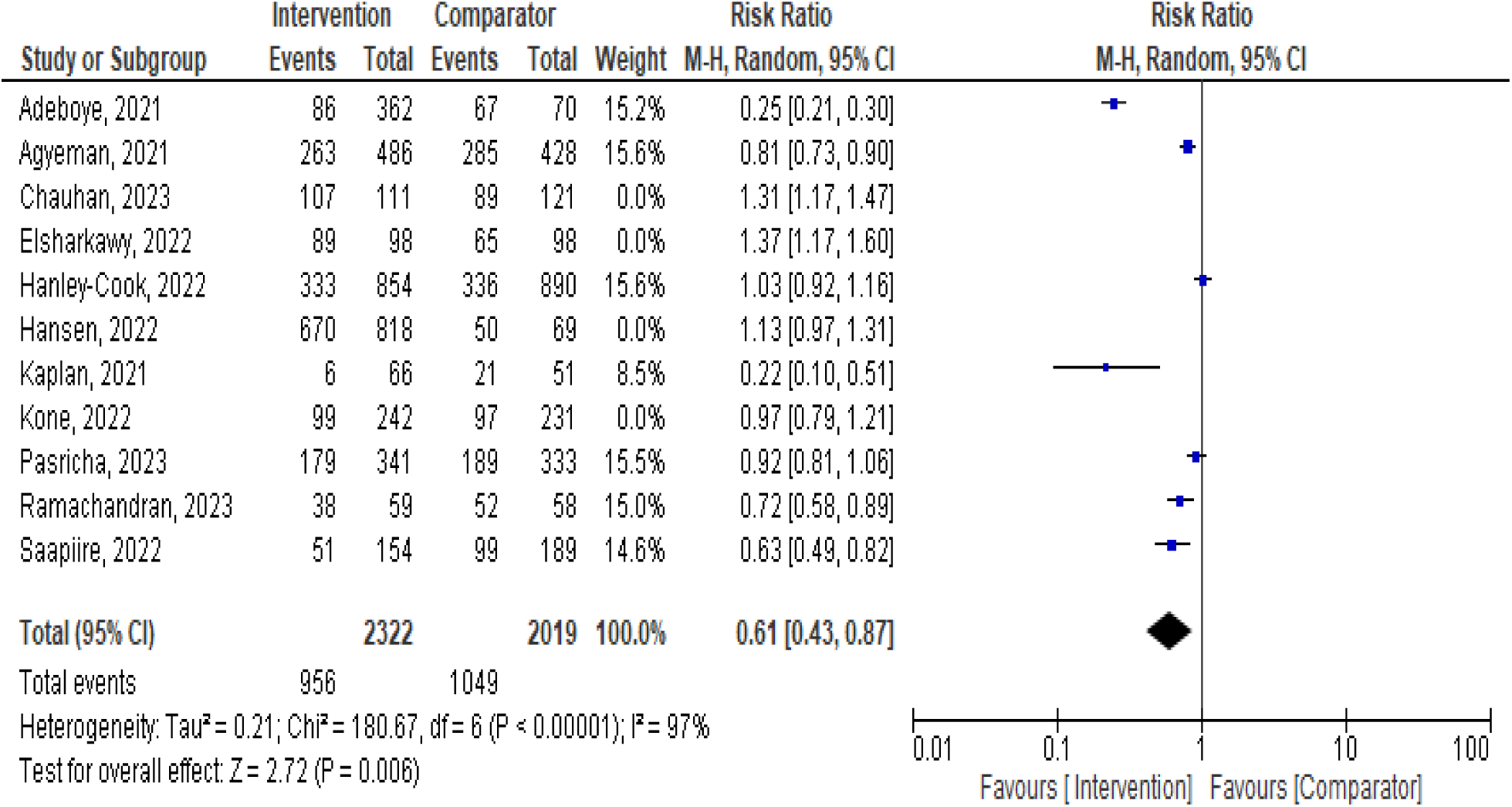
A forest plot of meta-analysis on the effect of maternal anemia interventions post-sensitivity analysis

**Figure 4.**
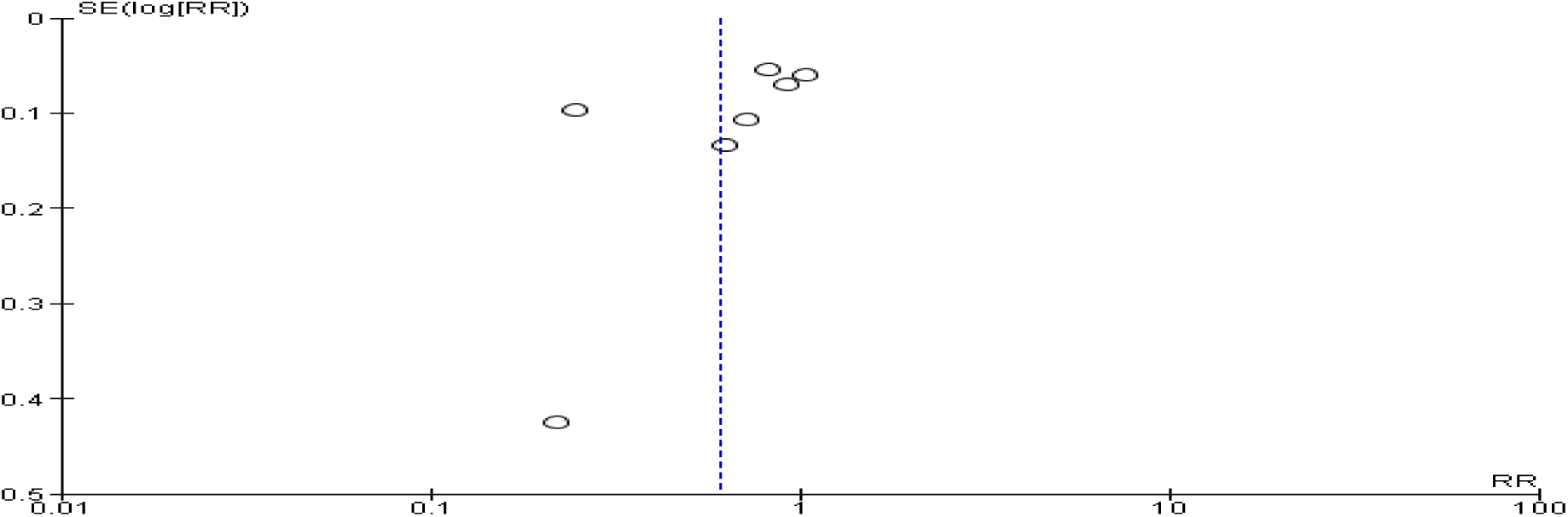
Funnel plot evaluating publication bias after sensitivity analysis. RR: relative risk.

### Subgroup Analysis and Investigation of Heterogeneity

Heterogeneity in the pooled effect estimates was considerably high for all 11 studies, contributed by 1,788 out of 6,129 (29 %) evaluated subjects, and thus it was necessary to perform subgroup analyses to identify possible variables or characteristics moderating the results obtained.

Subgroup analysis with random effects model was performed according to the type or form of the intervention used including dietary iron supplementation (n=2176), education or dietary information (n=786), I.V Ferric Carboxy-maltose (n=791), medicinal /herbal administration (n=914), I.V Sucrose (n=232) and other forms (n=1230). This showed still, a considerable heterogeneity (χ^2^5=38.92, P<.00001; I^2^=87.2%). The test for the overall effect for dietary iron supplementation (Z = 0.92 (P = 0.36), education or dietary information (Z = 0.05 (P = 0.96), Ferric Carboxy-maltose (Z = 1.01 (P = 0.31), and other interventions (Z = 0.51 (P = 0.61) all with substantial heterogeneity (I ^2^ > 90%) showed no significance difference (P < 0.05). Intravenous sucrose (RR 1.31, 95% CI 1.17, 1.47) (P = (Z = 4.70 (P < 0.00001) demonstrated poor prevention and management of maternal anemia at the gain of its comparator with 31 % while, the medicinal / herbal administration had a 19 % effect on prevention and management of maternal anemia (random-effects model RR 0.81, 95% CI 0.73, 0.90; *P* = 0.006) (*Figure 5*). The publication bias was further demonstrated by funnel plot (Figure 6).

**Figure 5.**
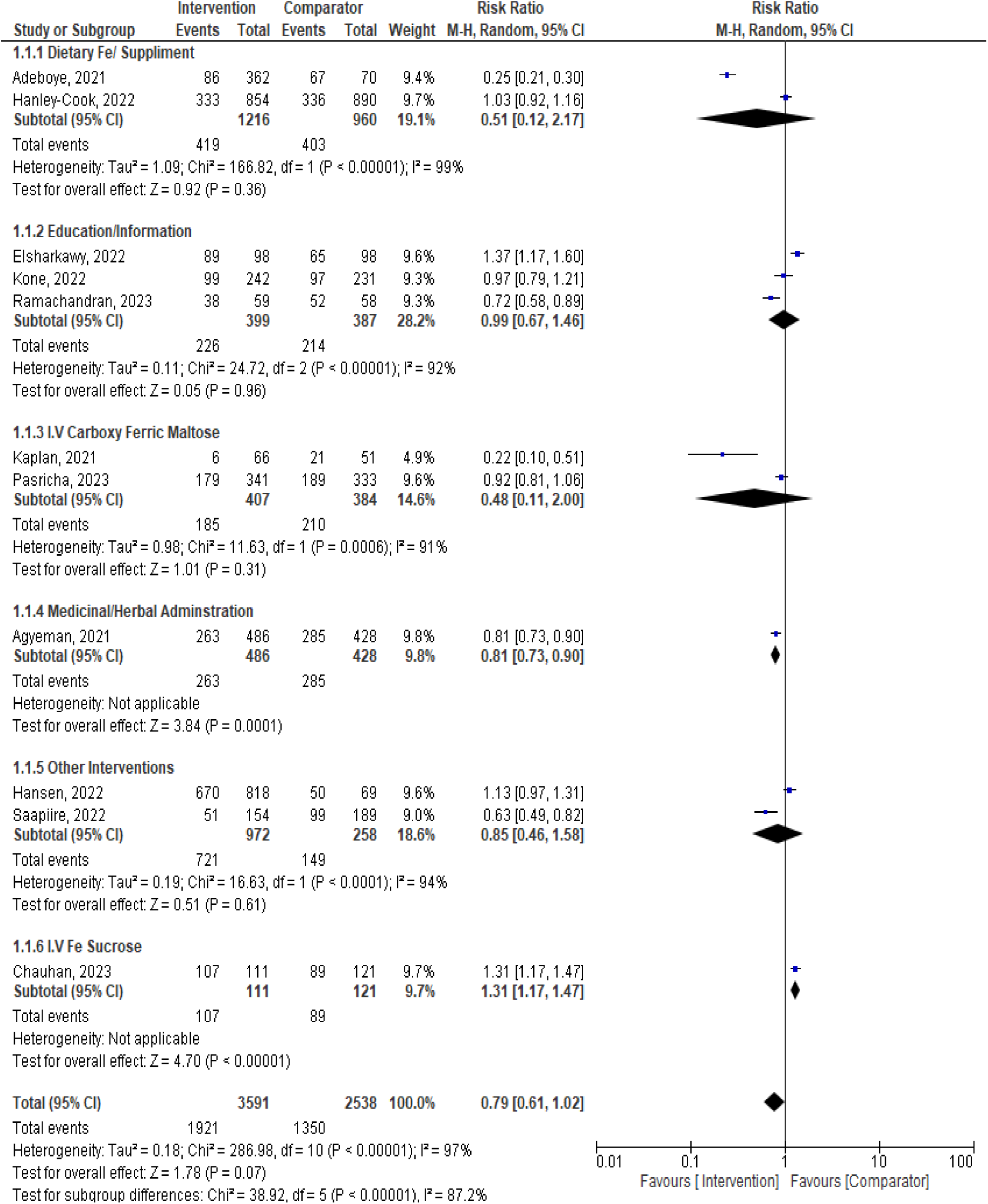
Subgroup analysis according the type or form of intervention showing similarly high heterogeneity as the full meta-analysis.

**Figure 6.**
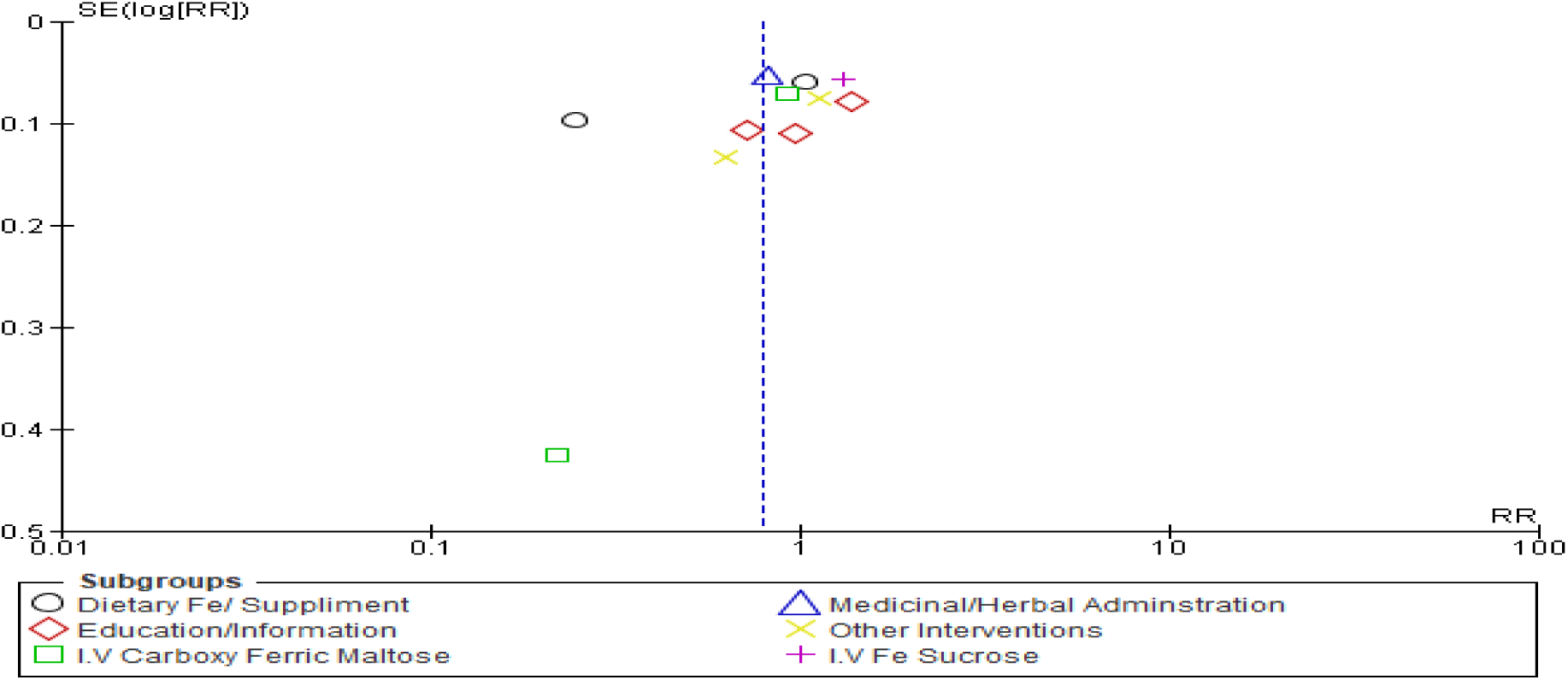
Funnel plot of the subgroup analysis-type or form of intervention.

**Figure 5.**
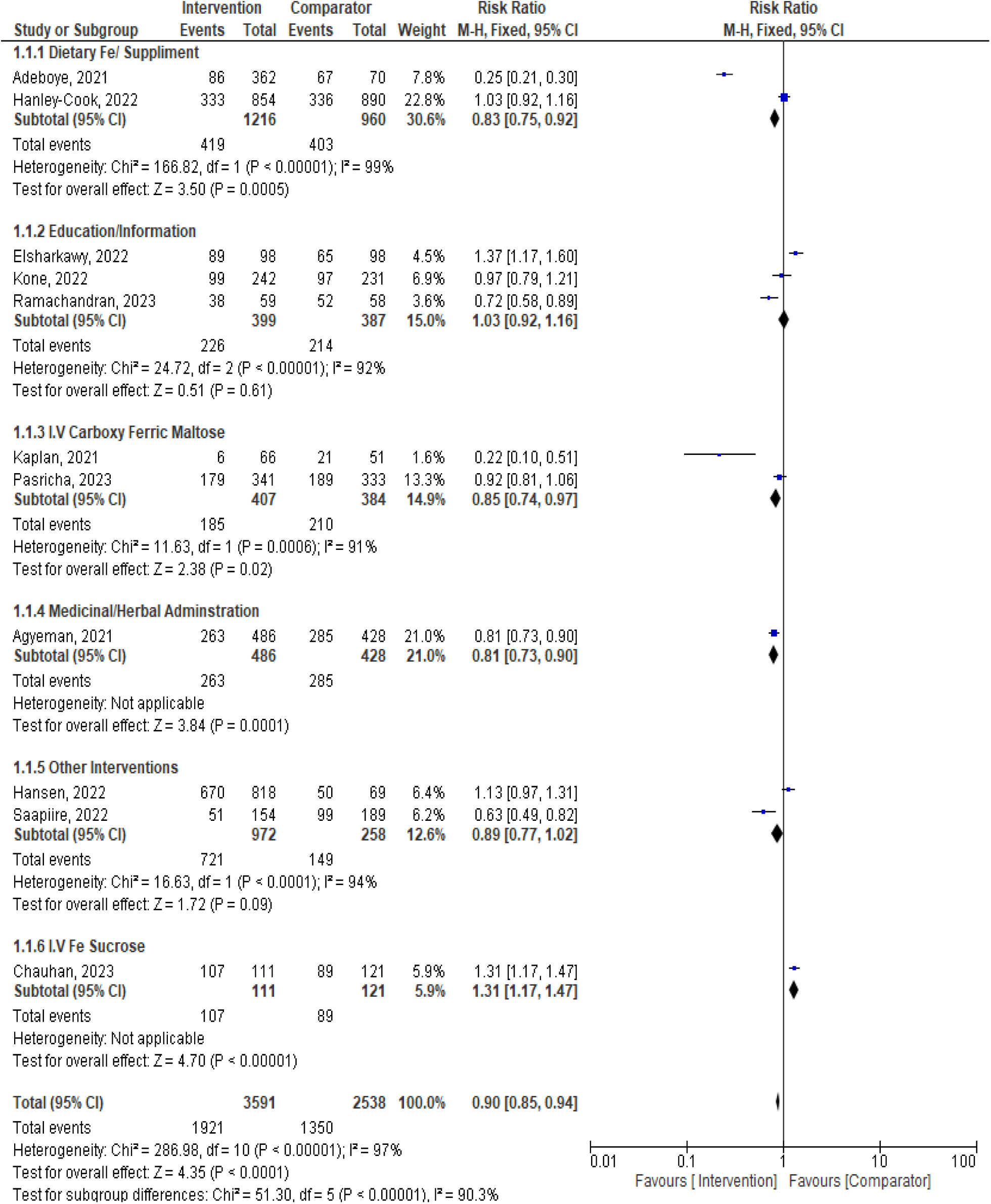
Subgroup analysis (fixed-effect model) according the type or form of intervention.

With fixed effect model, assuming that, one true effect size under lied each specific intervention form or approach, the sub-group analysis demonstrated that; dietary iron supplementation (RR 0.83 95% CI 0.75, 0.92), I.V Ferric Carboxy-maltose (RR 0.85 95% CI 0.74, 0.97) and medicinal/herbal administration (RR 0.81 95% CI 0.73, 0.90) interventions significantly influenced the prevention and or management of maternal anemia (P < 0.05), however, all still with high heterogeneity (I^2^ > 90%) (Figure 7).

**Figure 7.**
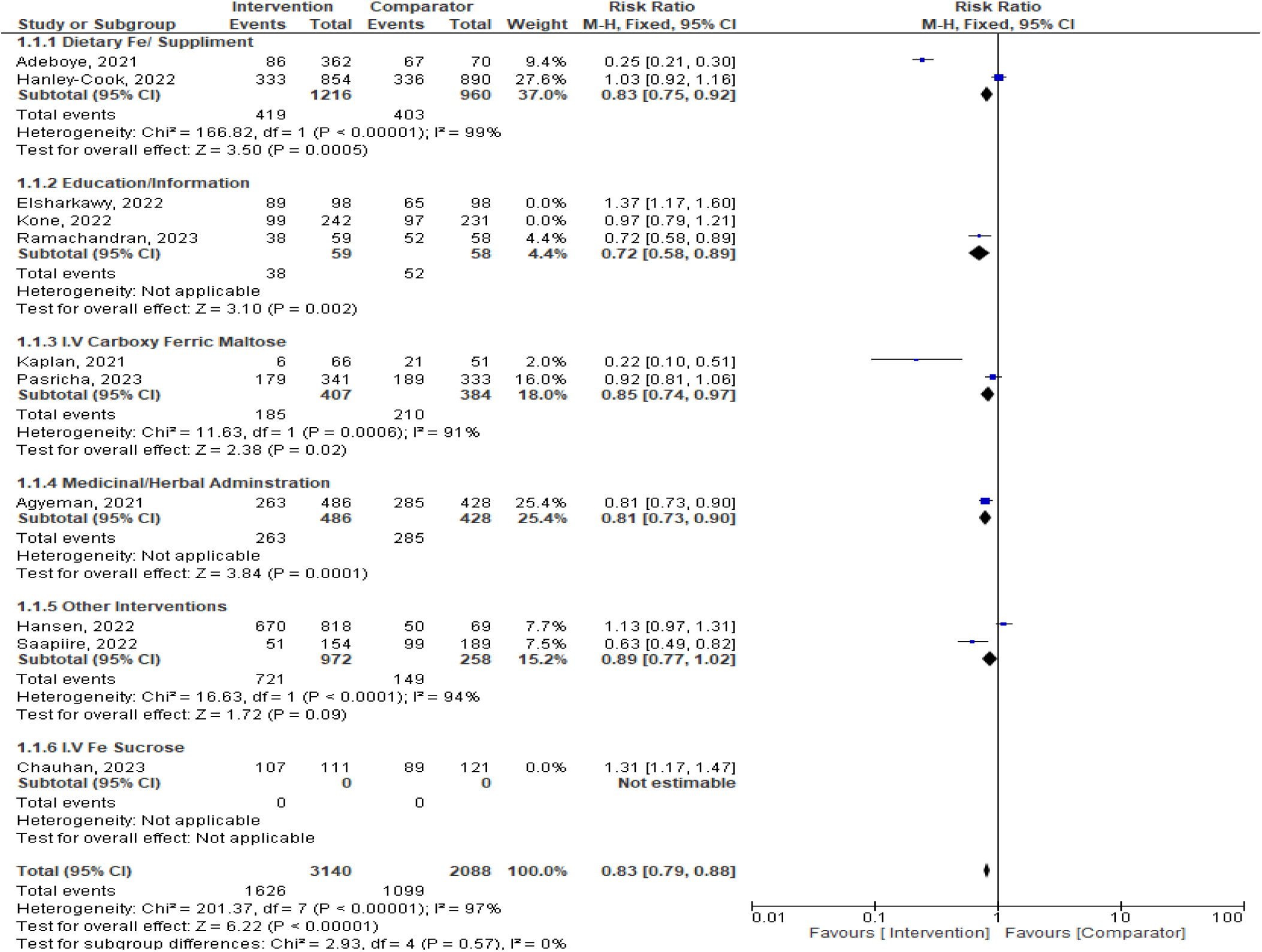
Sensitivity analysis on intervention type subgroups showing very low heterogeneity.

The high heterogeneity obtained from this prompted further, sensitivity analysis on each subgroup to ascertain the group that was most strongly associated with heterogeneity. Following this analysis on sub groups (n= 5228), by elimination of studies that caused the major heterogeneity[22], [23], [25], all the intervention approaches against maternal anemia showed a pooled positive effect of 17% ((fixed-effect model RR 0.83, 95% CI 0.79-0.88; P<.00001) (χ^2^4=2.93, P=0.57; I^2^=0%). Education or information given to pregnant women (n=117) showed a 28% effect (RR 0.72 95% CI 0.58, 0.89), medicinal /herbal administration 19% (RR 0.81 95% CI 0.73, 0.90) (n=914), iron supplementation 17% (RR 0.83 95% CI 0.75, 0.92) (n=2176) and I.V Ferric Carboxy-maltose 15% (RR 0.85 95% CI 0.74, 0.97) (n=791) (*Figure 7*), with greatly reduced publication bias and heterogeneity between the subgroups (I^2^ = 0%) shown by the funnel plot (*Figure 8*)

**Figure 8.**
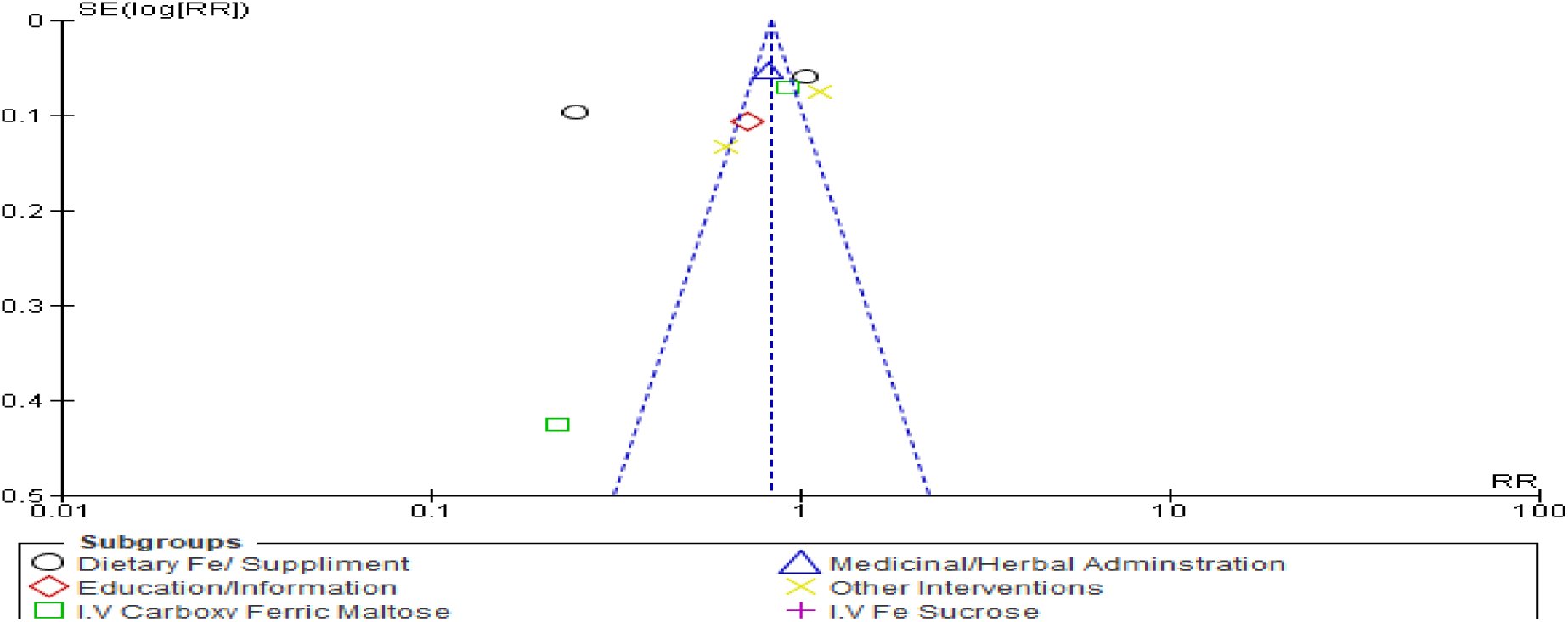
Funnel plot of the subgroup sensitivity analysis-type of intervention.

### Subgroup and sensitivity analysis on the possible covariates

#### a) Location of the pregnant women

Generally, maternal anemia interventions in the advent of COVID-19 demonstrated a higher (16%) and significant effect in African region (n=4580) as compared to Asia and European regions (fixed-effects model RR 0.84, 95% CI 0.79-0.89; P<.001) (χ^2^5=176.53, P<.00001; I^2^=97%) (*Figure 9*).

**Figure 9.**
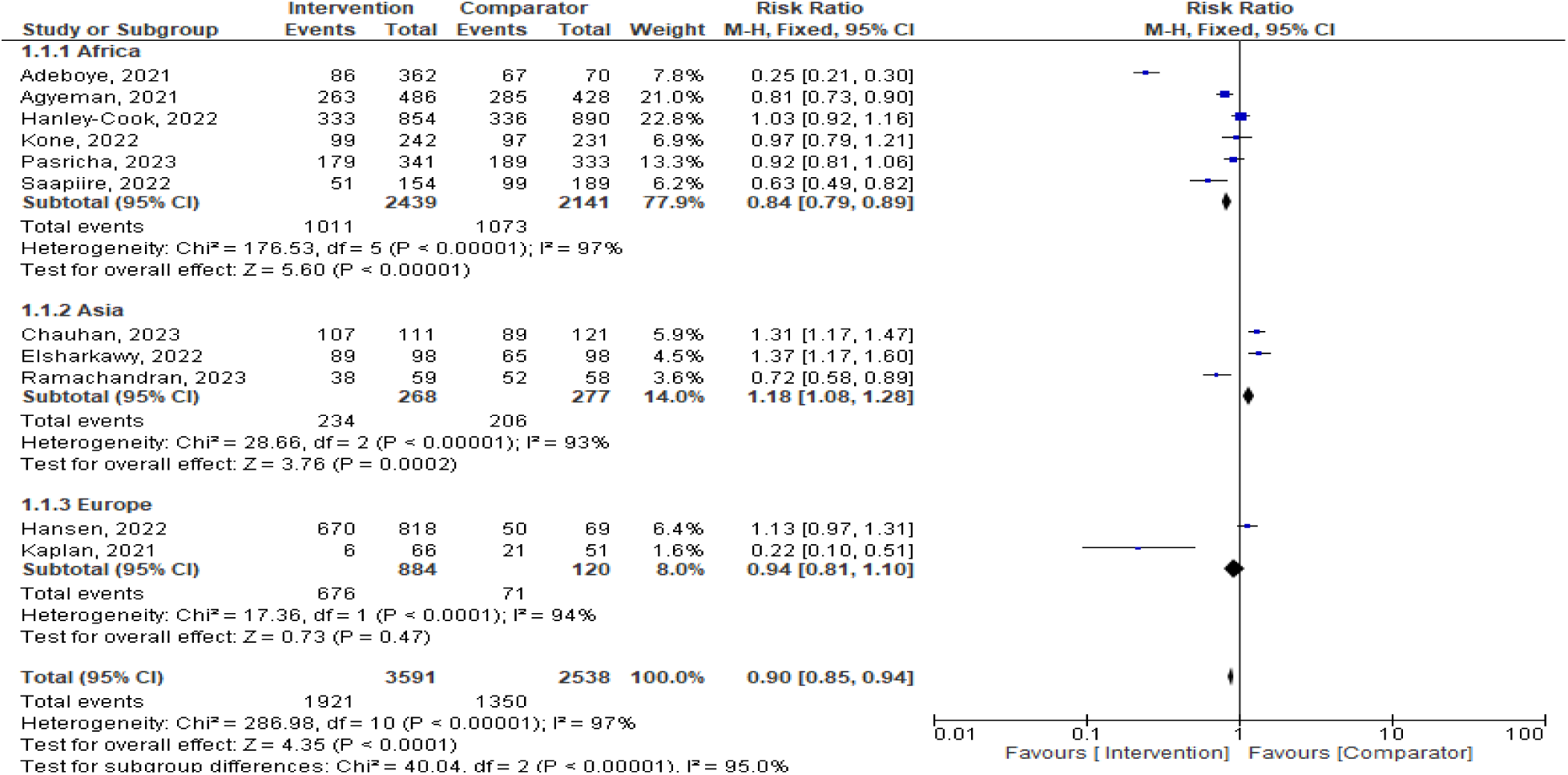
Subgroup analysis by the location of pregnant women in the advent of COVID-19.

#### b) Study setting

Similarly, multiple center studies (n=4580) showed a more significant predictive effect (16%) on maternal anemia intervention as compared to single center studies (n=1549) (fixed-effects model RR 0.84, 95% CI 0.79-0.89; P<.00001) (χ25=176.53, P<.00001; I^2^=97%) (*Figure 10*), with funnel plot clearly demonstrating that, most of studies close to the mean effect were multicenter associated with heterogeneity with only one study tending to signify homogeneity (*Figure 11*).

**Figure 10.**
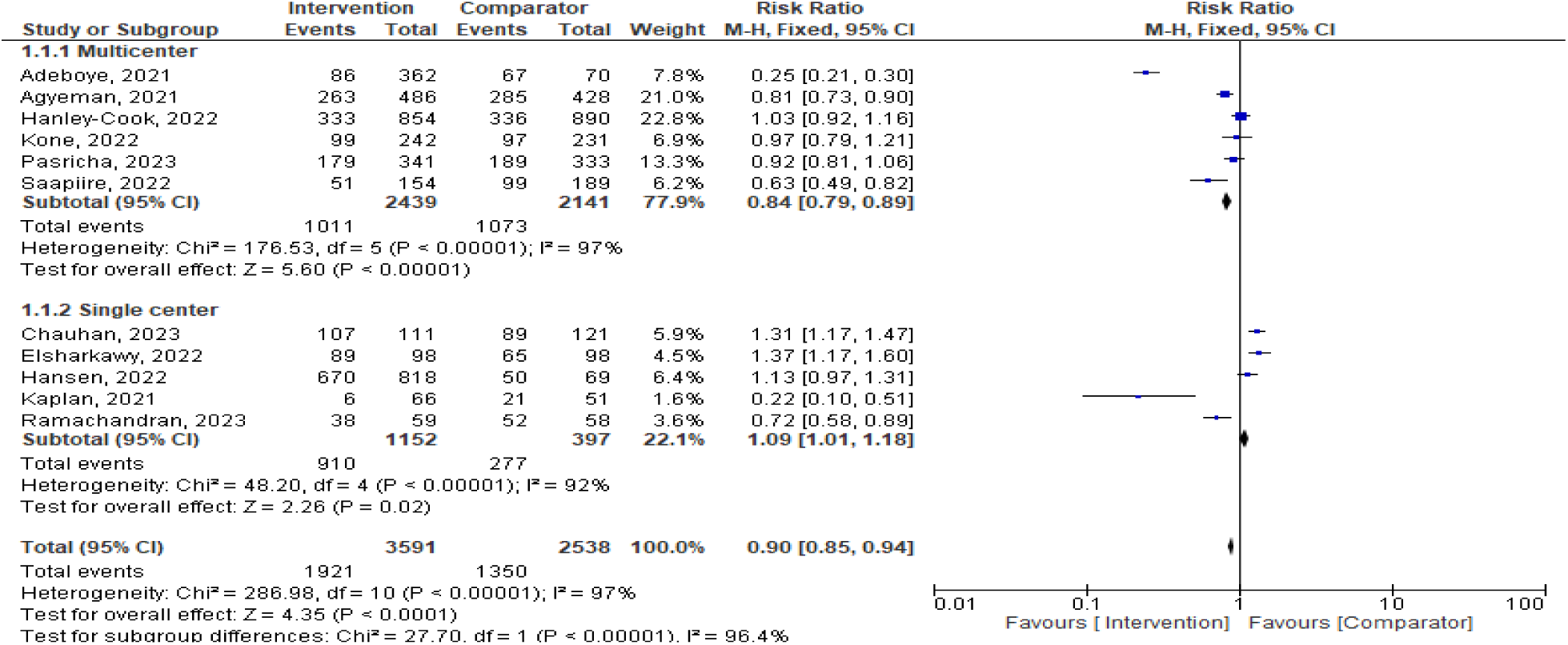
Subgroup analysis by the study setting

**Figure 11.**
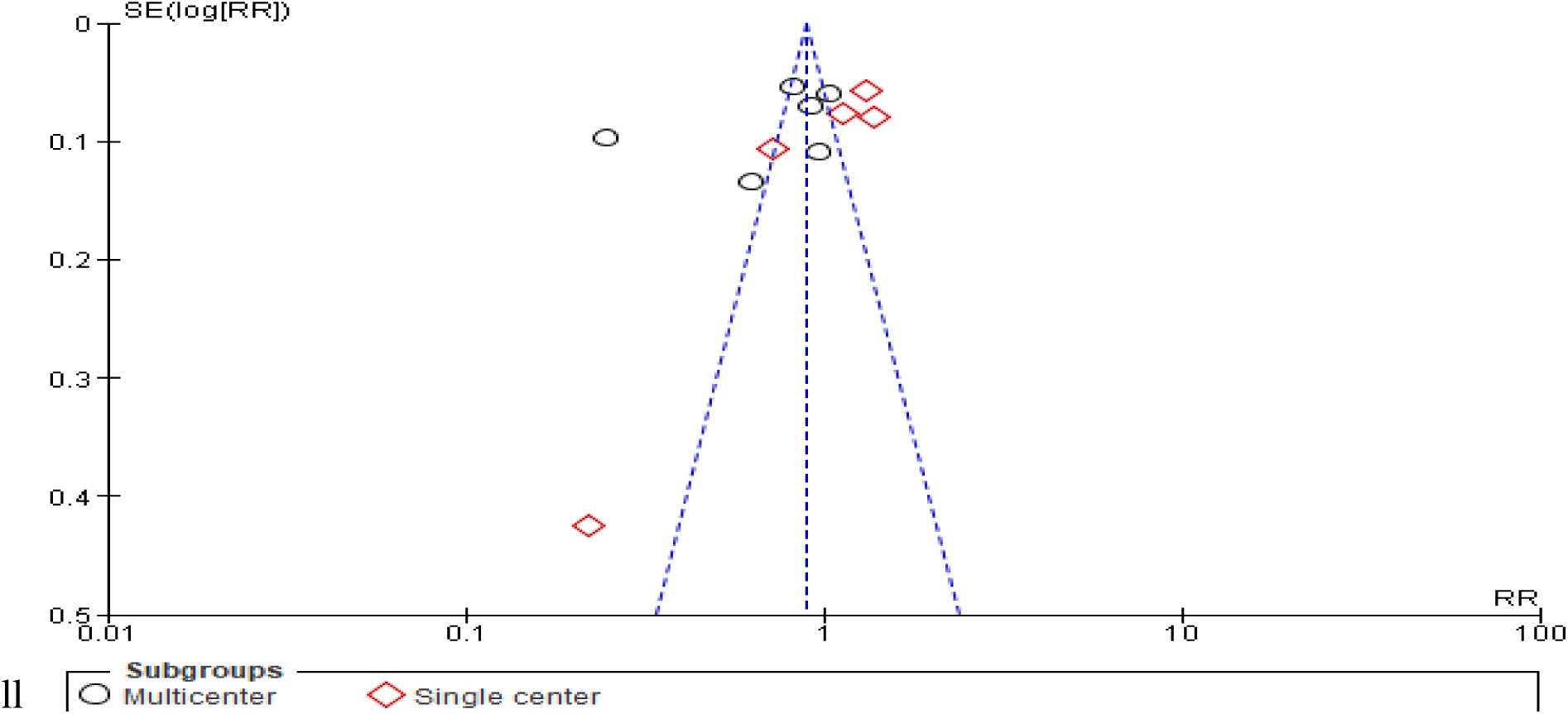
A funnel plot of subgroup analysis by the study setting showing single center studies (diamond shape) signifying higher heterogeneity

#### c) Time or year of data collection

Studies whose data were collected in the year 2020 in the advent of COVID-19 (n=2350) showed a more significant predictive effect (50%) on maternal anemia intervention as compared to other time or years of data collections (random-effects model RR 0.50, 95% CI 0.26-0.99; P<.05) (χ^2^3=167.34, P<.00001; I^2^=98%) (*Figure 12*). This was further evidenced with fixed effect analysis where, the year 2020 showed a 28% (random-effects model RR 0.72, 95% CI 0.67-0.78; P<.00001) (χ^2^3=167.34, P<.00001; I^2^=98%) (*Figure 13*).

**Figure 12:**
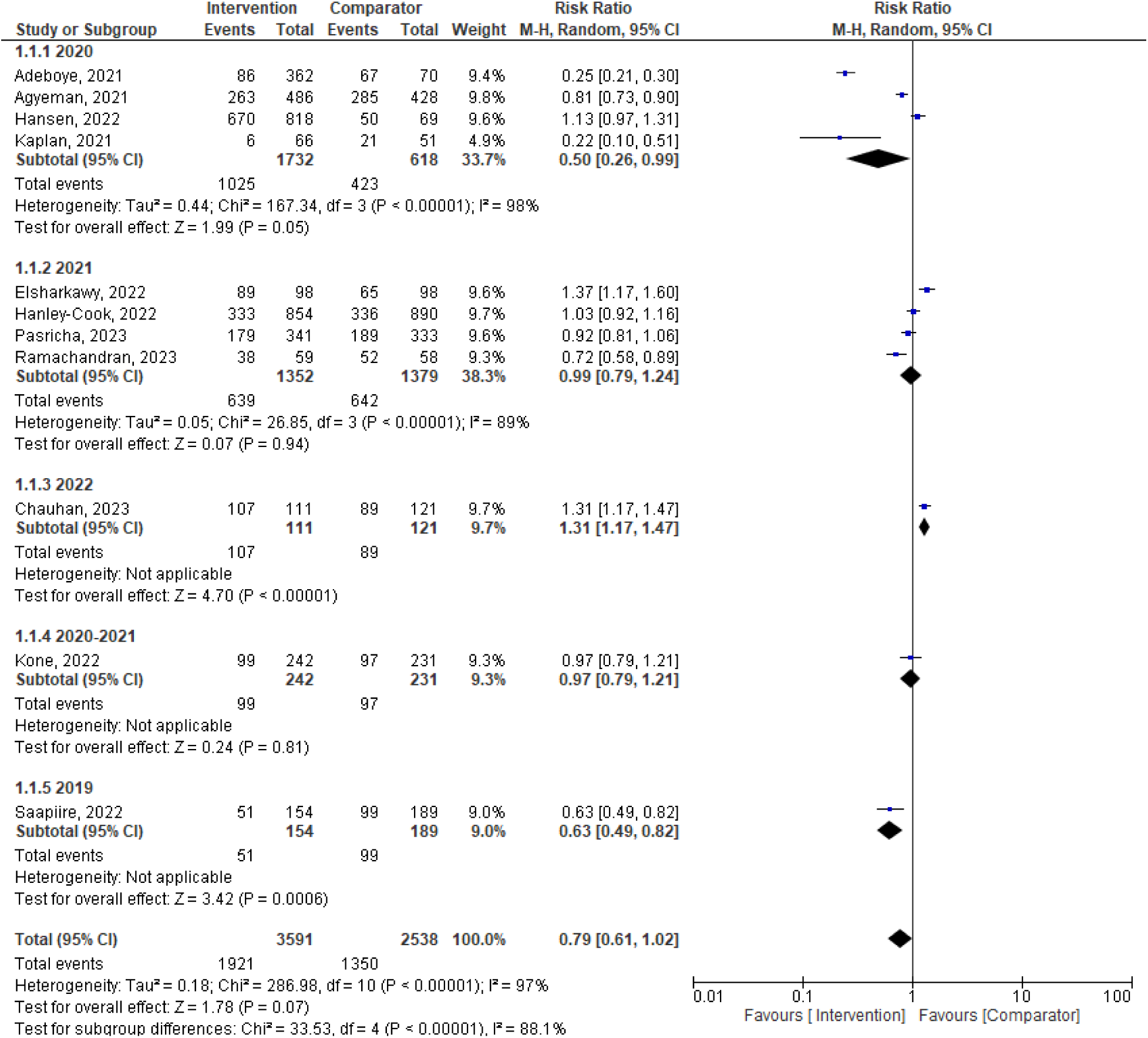
Subgroup analysis by the year the data was collected (Random Effects Model)

**Figure 13:**
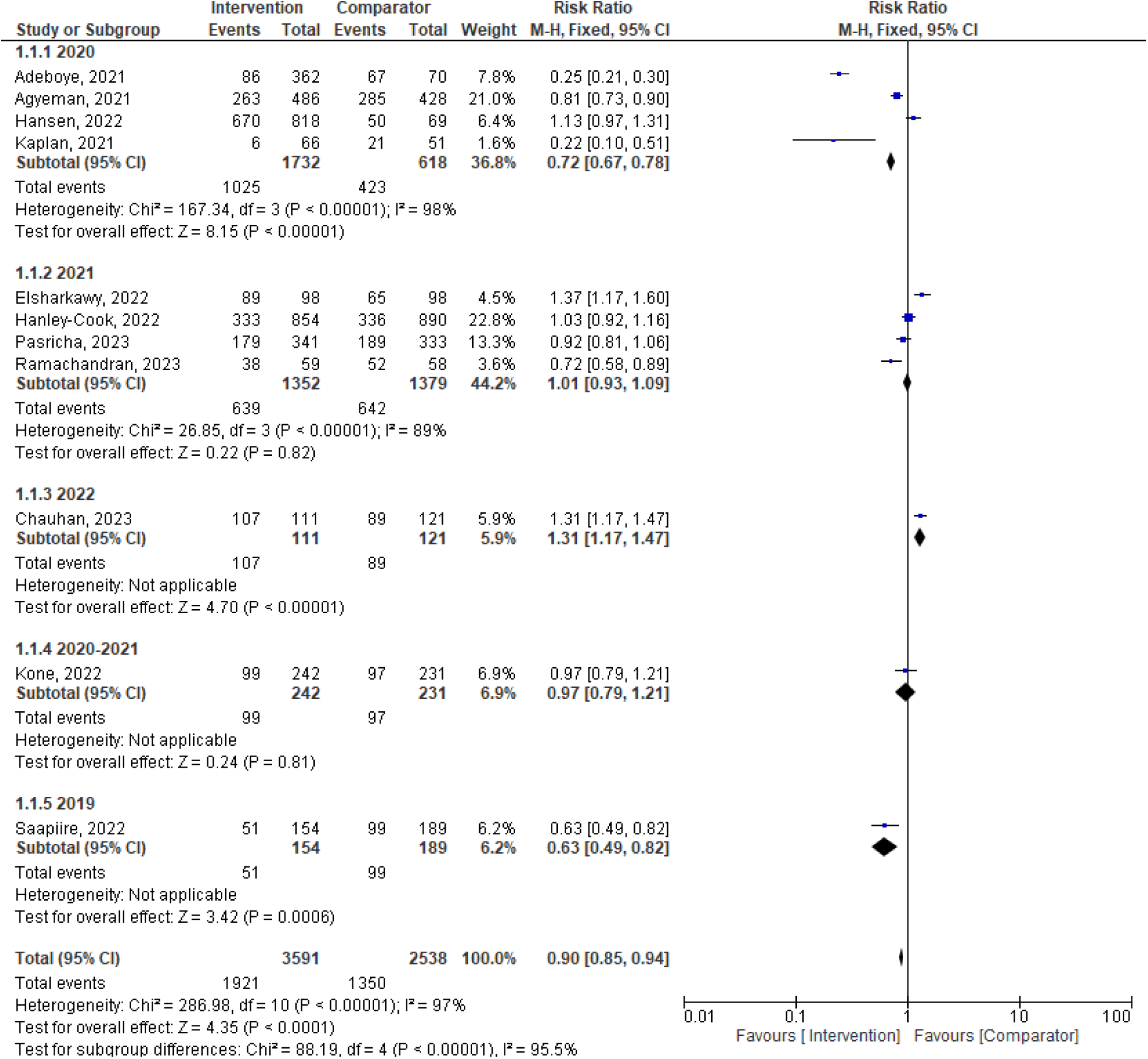
Subgroup analysis by the year the data was collected (Fixed-effect Model)

## Discussion

This review established that maternal anemia interventions in the advent of COVID-19 did actually contribute to control, prevention, treatment and or management of maternal anemia indicated by several parameters including hemoglobin level changes, mitigated clinical aspects associated with anemia, decreased need for postpartum blood transfusion, improved hemoglobin and plasma ferritin levels and significant reduction of anemia prevalence. Heterogeneity analysis revealed that the pooled effect estimate was considerably high considering all 11 included studies, contributed by 89% of the total patients evaluated. Further, sub-group and sensitivity analysis on each subgroup indicated a significant non-homogeneity.

This meta-analysis included 11 studies and revealed that the pooled intervention approaches had an effect on mitigating or reducing maternal anemia in the advent of COVID-19 by 39%. Previous research has indicated a similar range of effect during the pandemic based on iron supplementation and IFA interventions 1.39 (0.33, 2.45); P = 0.01* and 0.72 (0.36, 1.07); P < 0.00001 respectively [30]. The general net cumulative effect of the interventions on maternal anemia ranged from 23% to 81% [31]–[33]. This is implicated in past recent studies Additionally, another meta-analysis reported the outcome of interventions on maternal anemia during the COVID-19 pandemic[30]. This analysis adds to the extensive consensus in the literature, which should motivate further research investigating the key aspects inherent to anemia control in pregnancy during similar pandemics.

Prior studies have reported results that contrast with those presented here, with a better effect based on percentage reduction and or hemoglobin mean standard deviation change on controlling maternal anemia[9], [34], [35], however, not in the advent of COVID-19 pandemic. In addition, a meta-analysis that targeted only RCTs[36] showed superiority in preventing anemia by the intervention as compared with the control. Moreover, a study focusing on hemoglobin mean level change, demonstrated similar trend on improving anemia control in pregnancy[37]. Of concern, most studies as mentioned above have shown mixed outcomes relative to the outcome measure, with timelines of data collection in some being outside the scope as for the current study which has focused on the advent of COVID-19 pandemic. Further, the present systematic meta-analysis offers a more detailed view as it covers 11 studies from diverse regions capturing both single and multiple centers. The heterogeneity was high even after subgroup analysis adjustments as per the specific cluster of intervention.

However, with fixed model analysis, dietary iron supplementation (17%) I.V Ferric Carboxy-maltose (15%) and medicinal /herbal administration (19%) interventions significantly influenced the prevention and or management of maternal anemia, These findings are implicated in e-Library of Evidence for Nutrition Actions where, daily iron and folic acid supplementation during pregnancy improves anemic condition [38], while, efficacy and safety of intravenous ferric carboxy-maltose has been shown with similar conclusions [39]–[43] and on herbal medicinal remedies, past studies have also shown a similar trends of significant control of maternal anemia [44]–[46]. Of paramount importance to note in this regard is the fact that, the current study effect demonstrated by the pooled and specific interventions is seemingly lower as compared to the one demonstrated by the past studies mentioned here in. This can possibly therefore, be attributed to the influence of the COVID-19 pandemic compromising the different anemia interventions’ effectiveness.

The sensitivity analysis with fixed effect model on the subgroups by intervention type, further showed a pooled positive effect of 17%. Notably, education or information intervention to pregnant women showed a 28% effect in addition to medicinal /herbal administration), iron supplementation and I.V Ferric Carboxy-maltose. The greatly reduced publication bias and heterogeneity between the subgroups following this sensitivity analysis is an evidence that, education and information package to control maternal anemia is generally an efficient approach. Similar finding has been reported in a past study that, individual education through a pictorial handbook on anemia in conjunction with the counseling intervention program had a positive impact on hemoglobin and hematocrit levels for anemic pregnant women in their third trimester of pregnancy[47]. Mean change of hemoglobin levels were also found to be very significant in another study that established educational interventions can increase family support for maternal behavior in preventing pregnancy anemia such as improving adherence to taking iron supplements and high intake of food containing iron[35]. This fact on education. Information package intervention however, was better than the current findings in the advent of COVID-19 pandemic. Generally, education package on maternal anemia control is anchored under the integrated approach where all the other intervention methods are included as part of the package[48]. This reason may be the contributor of the current findings showing this cluster of intervention with the highest effect on maternal anemia control

Anemia intervention in African region generally recorded the highest effect as compared to other regions. This may not be due to the best practices though, as the prevalence of maternal anemia is higher in Sub-Saharan Africa[3], [49]–[51], however, this fact may be supported by the more interventions used to control anemia in Africa. Data collected from multiple center studies showed a more predictive effect (16%) of maternal anemia intervention as compared to single center studies Similar findings exist from other similar reviews although not during the COVID-19 pandemic[30], [52], [53]. In this context, the single center studies signified a major heterogeneity as compared to multiple center studies. Studies whose data were collected in the year 2020 in the advent of COVID-19 had a more significant predictive effect (50%) on maternal anemia intervention as compared to other time or years of data collections whose data after sub-analysis showed that, the trend on maternal anemia effectiveness decreased with time from the year 2020, to 2021 and 2022. This fact is clearly supported by a report asserting the availability of nutritious foods in particular is affected by COVID19 measures [54]. This was expected as nations concentrated on COVID 19 mitigation when it clearly became a pandemic after 2020 onwards. In addition, micronutrient intervention programs were affected during COVID-19 including disruptions of up to 75% were reported in Antenatal Care Programs selected countries during the first months of the lock-down [55] and stock-outs of IFA/MMS may have occurred with supply chains disrupted and programs no longer reporting stock information [56].

The effect and impact of specific disasters and or calamities on maternal anemia interventions has been investigated previously. In one study, the COVID-19 crisis exacerbated maternal and child undernutrition and child mortality in low- and middle-income countries[57]. Further, measuring the effects of COVID-19 disruptions on the delivery of essential health and nutrition interventions has proven challenging, as resilient, real-time information systems were not well established in many countries before the crisis [58]. A world Health Organization survey demonstrated an extent of disruptions across all healthcare services which solemnly may have led to disruption on the pregnancy anemia management and interventions[59].Adding on the same, an African based study found that, healthcare services utilization in the advent of COVID-19 were disrupted [60]. This could be expected to somehow affect and compromise the measures of mitigating the maternal. Another study demonstrated that COVID-19 pandemic has taken the world by storm affecting maternal health both directly and indirectly with complications of poor birth outcomes and maternal health[61]. This can be used to explain the reduced effect of maternal anemia interventions.

Resaerch shows that, in 2019, global anaemia prevalence was 29.9% (95% uncertainty interval (UI) 27.0%, 32.8%) in women of reproductive age, equivalent to over half a billion women aged 15-49 years. Prevalence was 29.6% (95% UI 26.6%, 32.5%) in non-pregnant women of reproductive age, and 36.5% (95% UI 34.0%, 39.1%) in pregnant women. Since 2000, the global prevalence of anaemia in women of reproductive age has been stagnant, while the prevalence of anaemia in pregnant women has decreased slightly [62]. Although more and more information is accumulating daily since COVID-19 pandemic, subjective factors on pregnancy and the effect of the pandemic on health systems in African nations may have compromised the right trajectory towards addressing anemia in general[63]. Given this, a couple of interlinked factors including any similar pandemic should be considered together as a single risk factor for maternal anemia.

Several factors limit the interpretation of the present study. First, the vast majority of studies included in the meta-analysis were retrospective epidemiological studies with only four RCTs deemed more reliable in terms of evidence on ascertaining the effect an intervention on an outcome. Second, some of the included studies did not distinguish the age range of the participants as well as the stage of the gestation period. Third, COVID-19 may have also indirectly contributed to the anemic condition measures by hemoglobin levels among the pregnant women. Future studies may respond to these issues by defining clearly, the scope of direct and indirect effect of a similar pandemic of disaster for better prompt mitigation measures.

Multiple factors are responsible for recurrent pregnancy loss, although any other phenomena such as unforeseen pandemic may compromise anemia control in this vulnerable population. Similarly, different interventions for maternal anemia may be differently compromised. Thus, pregnant women should be screened to assess the best option for an intervention and further, outline in advance, the feasible approaches that may aid the best effect achievability in any form of a pandemic or disaster.

Finally, the effect of the COVID-19 pandemic on anemia interventions in pregnancy should also be further examined and clarified. In addition, key healthcare stakeholders and experts should pay more attention to the perceived changes associated with any phenomena that is deemed to be a risk to the positive outcome of maternal anemia management and control. Further research may aim to determine the mechanisms that drive or decrease this risk of compromised maternal anemia interventions either directly and indirectly.

This meta-analysis revealed that in the advent of COVID-19, different measures of mitigating maternal anemia were compromised. This was demonstrated by a low effectiveness trend from the year 2020 to the year 2022. Although even before the COVID-19 pandemic there has been some perceived challenges on the same, during this period, even the most effective and recommended interventions against maternal anemia were somehow affected in the advent of COVID-19.The SARS-CoV-2 or any future similar epidemic should serve as an impetus for further research on maternal anemia and the effectiveness of the key recommended interventions, and to map out the most feasible intervention approaches as well as proximate factors associated with the reduced effectiveness of these interventions for continued improvement.

## Data Availability

All data produced in the present study are available upon reasonable request to the authors

## Notes

### Competing Interest Statement

The authors have declared no competing interest.

### Funding Statement

This study did not receive any funding

